# SARS-CoV-2 antigenemia/viremia masks seroconversion in a COVID-19 patient

**DOI:** 10.1101/2021.01.26.21250561

**Authors:** Konstantinos Belogiannis, Venetia A. Florou, Paraskevi C. Fragkou, Stefanos Ferous, Loukas Chatzis, Aikaterini Polyzou, Nefeli Lagopati, Aikaterini Touliatou, Demetrios Vassilakos, Christos Kittas, Athanasios G. Tzioufas, Sotiris Tsiodras, Vassilis Gorgoulis

**Author notes:** **Corresponding author**: Prof. Sotirios Tsiodras, 4^th^ Department of Internal Medicine, Attikon University Hospital, Medical School, National and Kapodistrian University of Athens, 1Rimini Str, Chaidari, Attiki, 12462, Greece. Equal contribution as first authors (Konstantinos Belogiannis, Venetia A. Florou, Paraskevi C. Fragkou).

## Abstract

Immune responses against SARS-CoV-2 have been vigorously analyzed. It has been proposed that a subset of mild or asymptomatic cases with undetectable antibodies may clear the virus in a T-cell cytotoxic-dependent manner, albeit recent data revealed the importance of B-cells in that regard. We hypothesized that underdiagnosed antigenemia/viremia may conceal humoral response possibly through immunocomplex formation. We report the first case of late-onset seroconversion detected following decline in antigenemia/viremia levels. Consequently, classification of at least a subset of COVID-19 cases as non-responders might not represent a true immunobiological phenomenon, rather reflect antibody masking due to prolonged antigenemia/viremia.

## Introduction

Severe Acute Respiratory Syndrome Coronavirus 2 (SARS-CoV-2), the culprit of an ongoing pandemic, continues to engender detrimental effects on healthcare systems worldwide leading to serious socioeconomic consequences. Following an incubation period of 2-14 days infected individuals experience a heterogeneous clinical course of the so-called Coronavirus Disease-2019 (COVID-19), ranging from asymptomatic infection to critical illness^1,2^. Similarly, symptomatic infection comprises of a wide array of clinical manifestations from localized disease affecting preferentially the respiratory and occasionally the gastrointestinal tract, to multi-systemic organ involvement^3,4^.

The brisk induction of pro-inflammatory responses, the so-called cytokine storm syndrome, is currently regarded as the major contributor of COVID-19-related multi-organ dysfunction^5^. However, there is sparse evidence that contradicts this supposition, implying a possible underestimated degree of viral-induced organ cytotoxicity^6^. Although the route of viral dissemination to other organs is still a subject of debate, growing evidence suggests this occurs haemotogenously^4,5,7^. Nonetheless, SARS-CoV-2 viremia and antigenemia have only been documented in disproportionally low rates than expected and the clinical significance of their presence remains undetermined^8–11^.

Besides the role of viremia in COVID-19 pathogenesis, another ambiguous aspect of the disease is the host’s immune response against SARS-CoV-2, and more specifically, the diversity of the humoral response level among SARS-CoV-2-infected patients. Based on currently available serological assays, it is evident that the majority of COVID-19 patients seroconvert within 2 weeks post symptom onset (p.s.o), whereas delayed (beyond the 2^nd^week p.s.o) or even absent antibody responses (non-responders) have also been documented **(Figure 1A)**^12^. The latter is particularly true in asymptomatic or paucisymptomatic patients^12^.

**Figure 1:**
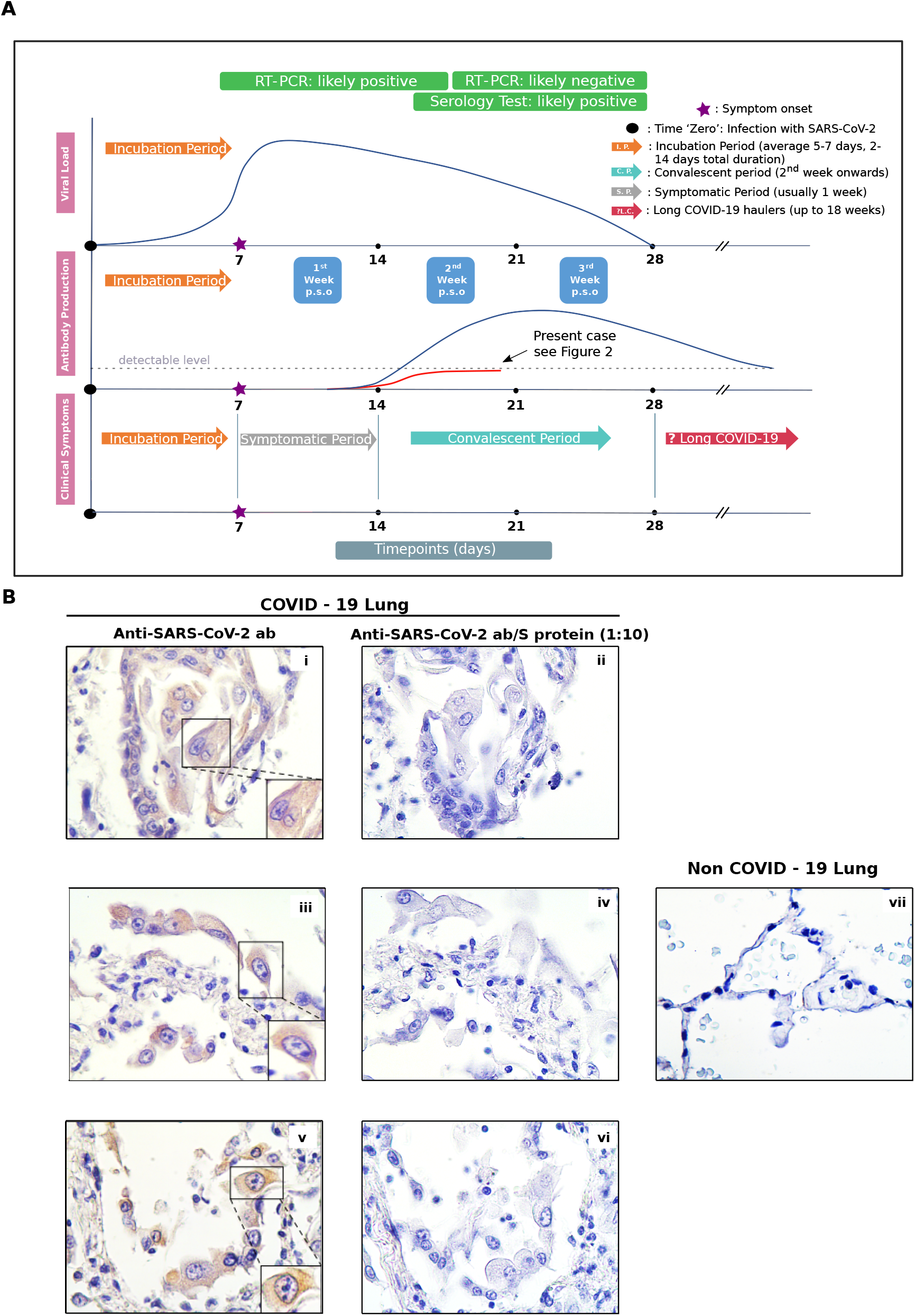
**A**. Antibody responses and viral load temporal kinetics as they correlate with clinical symptoms are depicted. Estimated time intervals are based on data from several published studies. The graph in red illustrates the antibody response pattern of our patient - who is considered as late/non-responder - relatively to that mounted in most COVID-19 individuals. **B**. Immunohistochemical images from lung tissues of a positive COVID-19 patient (images i-vi) and a negative control (image vii). Cytoplasmic staining of Type II Pneumocytes is seen following incubation with secondary Anti-SARS-CoV-2 monoclonal antibody (479-G2) with (ii, iv, vi) and without (i, iii, v) competition with excess (one in ten dilution) viral Spike (S) protein. Upon competition, cytoplasmic signal is lost (ii, iv, vi).

This divergent pattern of humoral response has also been observed in the other two beta- coronaviruses (SARS-CoV and Middle East respiratory syndrome coronavirus, MERS-CoV) as well as other viral strains, such as human papillomavirus (HPV) and human rhinoviruses^12-14^. This phenomenon of undetectable antibody titers following convalescence constitutes a paradox which has neither been studied nor satisfactorily explained.

To explain this paradox in COVID-19, we sought to examine the hypothesis that the presence of the virus and/or viral fragments (viremia and/or antigenemia) in the serum may mask seroconversion. If this is true, then two conditions should be met: 1) viral RNA should be detected in the blood stream, and 2) if the absence of seroconversion is due to antigen/viral-mediated saturation of circulating antibodies (i.e immunocomplexes formation), then the progressive decrease of antigenemia/viremia should be followed by increasingly detectable antibody titers^15^. Herein, we report to the best of our knowledge, the first case as a proof-of-concept supporting the proposed scenario.

### Case presentation

We report the case of a young male in his 20’s who presented with a 24-hour history of fever up to 38.4°C, without any additional signs or symptoms (**Figure 2A**). Due to a recent history of close contact with a confirmed COVID-19 case, nasopharyngeal swab from the patient was obtained and tested for SARS-CoV-2 with reverse transcription polymerase chain reaction (RT-PCR) on day 2 p.s.o, confirming the diagnosis (Cycle threshold value (Ct)=15). Subsequently, the patient self-isolated at home for two weeks, as per national infection control protocols. His fever subsided within a few days. Besides antipyretics, the patient did not receive any other medications. However, upon completion of the two-weeks’ isolation, low grade fever (up to 37.4°C) recurred. A repeat RT-PCR test on day 15 p.s.o. was positive (Ct=30) (**Figure 2A**). Fever finally resolved 25 days after symptom onset.

**Figure 2:**
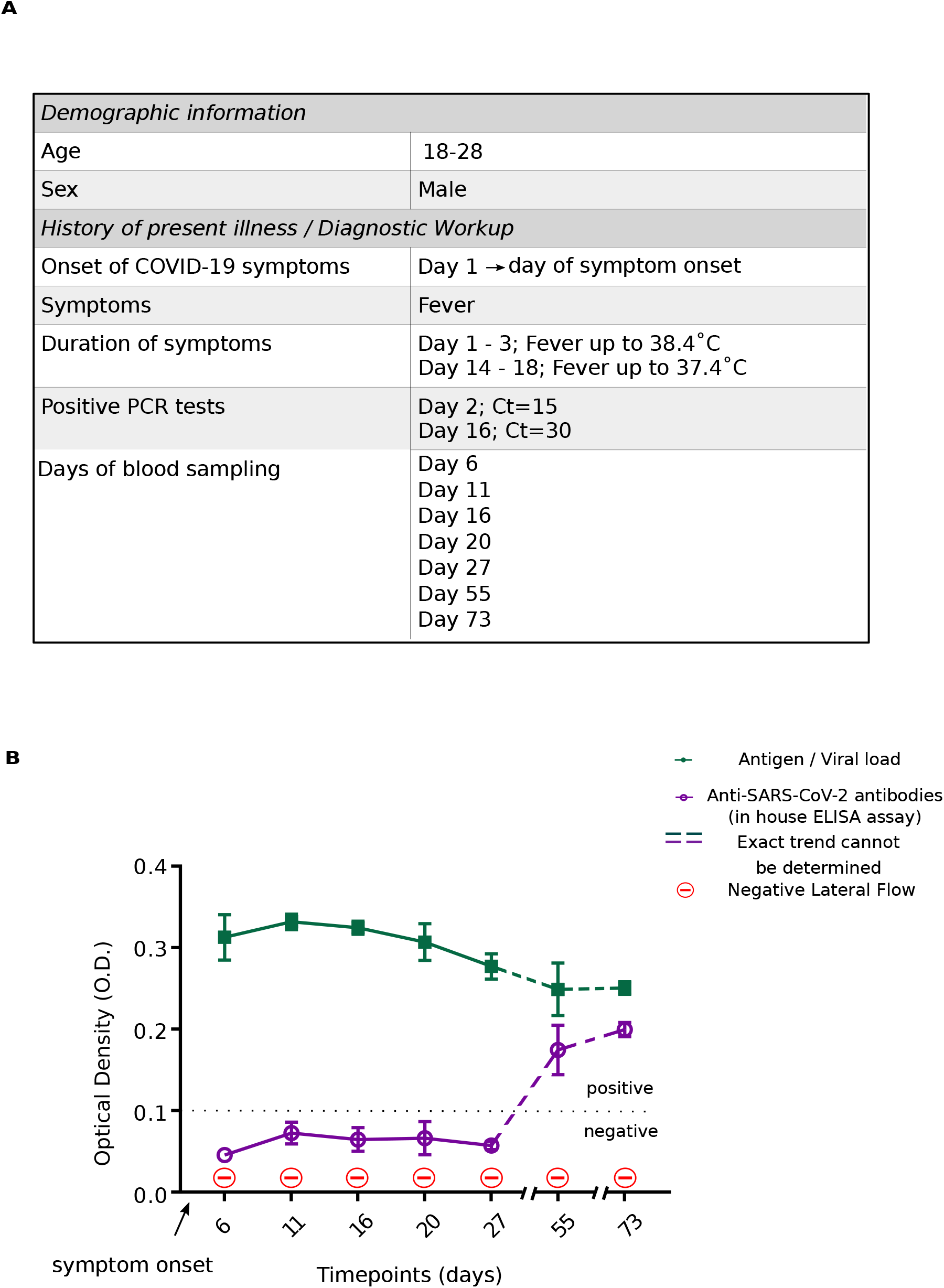
**A**. Table summarizing patient’s (presented case) clinical characteristics. **B**. Diagram depicting kinetics of antigen/viral load as relate to antibody generation detected by two separate immunodiagnostic assays (in house ELISA and Lateral Flow). Time-dependent decline in the antigen/viral load is associated with seroconversion 55-days post symptom onset in the case of our ‘in house’ ELISA, whilst it failed to occur according to Lateral Flow assay.

A series of serum samples for the detection of antibodies against SARS-CoV-2 were collected at regular intervals (every 4-5 days) starting on day 6 p.s.o up to day 27 p.s.o, followed by two additional samples on days 55 and 73 **(Figure 2A)**, a timeframe that exceeds the expected seroconversion window **(Figure 1A)**.

## Materials and Methods

### Monoclonal antibody production

The monoclonal antibodies 480-S2 and 479-G2 used in the present immunoassays have been generated by immunizing mice against the receptor binding domain (RBD) region of the spike (S) protein of SARS-CoV-2 via a modification of the method described by Koehler and Milstein^16^. Following vigorous immunosorbent assay selection cycles, clones exhibiting the required sensitivity, specificity and reproducibility were selected for downstream applications as detailed below. These have been extensively validated **(Figure 1B)** and are under proprietary rights (patent application no.:22-0003846810).

### Double antigen ELISA for antibody detection

High-binding plates precoated with 1.5μg/ml S-trimer (Trenzyme, GmBH, Germany) were blocked with 300μl of 4% Bovine Serum Albumin (BSA) following incubation for 1 hour at Room Temperature (RT). Then, 50μl of serum samples were loaded in duplicates (1:1 dilution) and incubated at 4°C overnight. Next, 50μL/well of S protein conjugated to Horseradish Peroxidase (S-HRP) (5:12000 dilution) were loaded and incubated for 45 minutes at RT. After appropriate washing, 50μL/well of 3,3’,5,5’-Tetramethylbenzidine (TMB) were added and let to incubate for 10 minutes at RT in the dark. Subsequently, 50μL of phosphoric acid were used for reaction termination and the absorption was quantified using a microplate reader at 450nm (cut-off value: 0.100). Washing was performed at appropriate steps using Phosphate Buffer Saline/0.1% Tween (PBSTx5). Following testing on 150 negative pre-COVID-19 and 250 RT-PCR positive samples, validation data revealed 90.5% sensitivity and 95% specificity **(Suppl. Figure 1i)**.

Our ‘in house’ method was cross-referenced to an FDA-approved and independently validated enzyme linked immunosorbent assay (ELISA) (Euroimmun, Luebeck, GmBH, Germany), by testing a panel of 321 anonymised samples (Pearson’s Chi-squared test with Yates’ continuity correction; p-value=0.5288)^17^. With regards to the presented case, the only discrepancy between the two ELISA methods was day 27 (2 weeks beyond the expected seroconversion window) where Euroimmun was positive and ours was negative. Moreover, antibody detection was tested by a commercially available Rapid Antigen Detection test, as well (lateral flow technology by Prognosis, BIO-SHIELD 2019-NCOV IGG, Catalog number: C1148/C1196).

### Sandwich ELISA for SARS-CoV-2 antigen detection

High-binding plates precoated with 2μg/ml of monoclonal antibody 480-S2 were blocked with 300μl of 4%BSA/0.05%Tween blocking buffer following incubation for 1 hour at 37°C. Subsequently, 50μl of serum samples (1:1 dilution) were loaded in duplicates and were allowed to incubate overnight at 4°C. Following adequate washes with PBST, 50μl of secondary antibody 479-G2 solution labelled with HRP (G2-HRP at a concentration of 1:50000) were loaded into each well, and after a 30min incubation period at RT in the dark, development of the signal was performed using 50μl of TMB substrate. Following a 10min incubation time, equal volume of phosphoric acid was introduced to terminate the reaction and signal was quantified as above (cut-off value: 0.100). Performance characteristics of the test were calculated following testing on 100 samples (28 RT-PCR positive and 72 negative pre-COVID-19 samples), revealing a sensitivity of 93% and specificity of 99% **(Suppl. Figure 1ii)**.

### Immunohistochemistry

Immunohistochemistry was performed using the anti-SARS-CoV-2 monoclonal antibody (479-G2). The Novolink Polymer Detection System (Leica Biosystems) was used for development of the signal and hematoxylin for counterstaining **(Figure 1B, Suppl. Figure 2B)**. The specificity of the immunohistochemical signal was confirmed by 1) omitting the primary antibody and 2) performing competition with the corresponding spike antigen **(Figure 1B)**.

### Molecular Detection of SARS-CoV-2 RNA

RT-PCR was implemented via a validated protocol as previously reported^18^.

### Next Generation Sequencing

Next Generation Sequencing (NGS) of the viral genome in the presented case was performed as detailed in Supplementary Materials and Methods and in **Suppl. Figure 3**.

**Figure 3:**
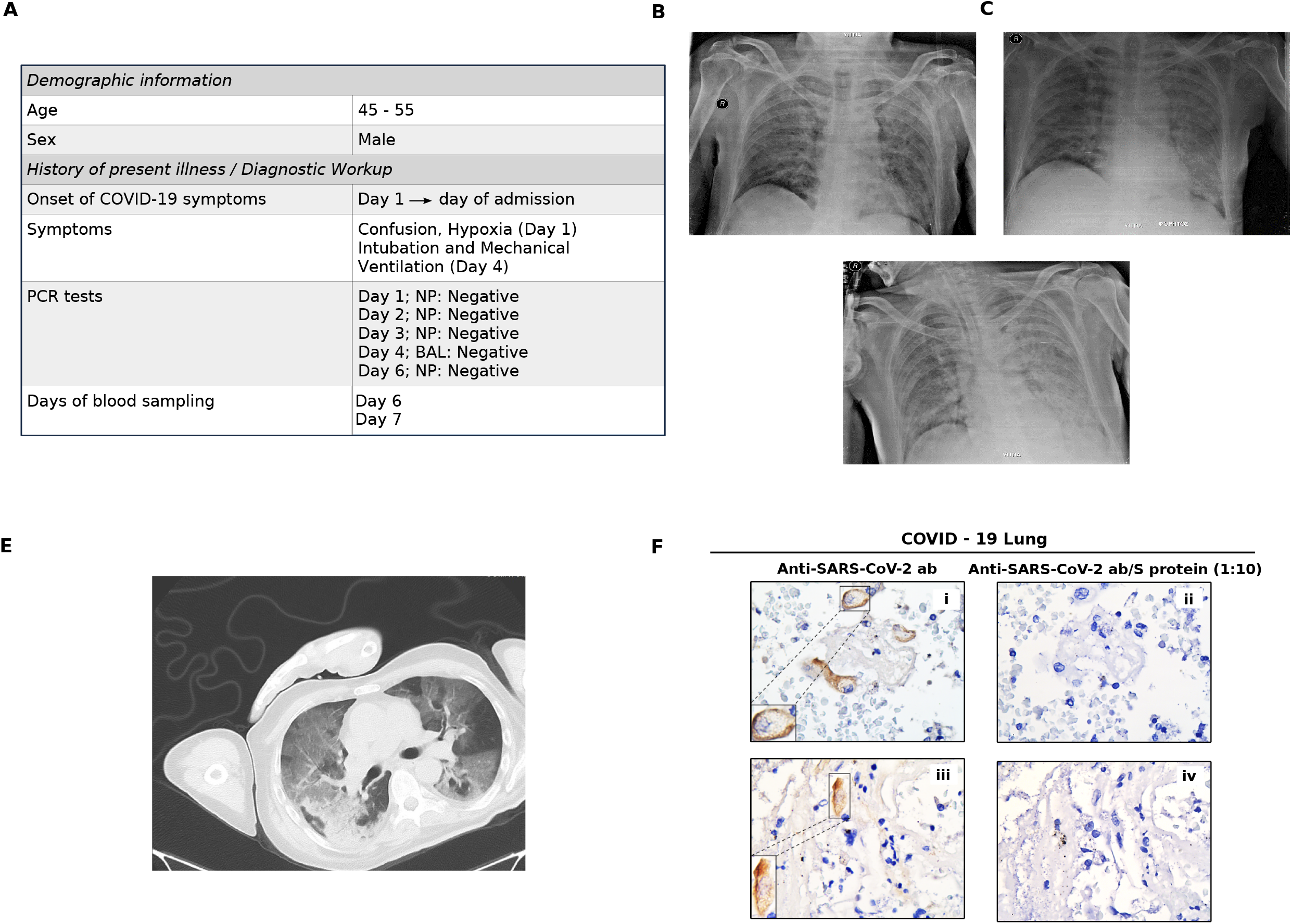
**A**. Table summarizing sampling dates and types of specimens obtained for diagnostic purposes in a separate patient with high clinical suspicion of COVID-19. **B, C, D:** X-RAYS obtained at different time points during patient’s clinical course (on admission, at clinical deterioration (Day 4) and post-intubation, respectively). Progressively worsening patchy infiltrations as well as air bronchogram can be visualized in affected lung fields. **E**. Computed Tomography (CT) of the lungs obtained at time of clinical deterioration (following Chest X-RAY; **Figure 2C**) demonstrating diffuse ground glass opacities in affected lung fields, typical for COVID-19. **F**. Immunohistochemical confirmation of SARS-CoV-2 infection in cadaveric lung tissue material derived from the same patient.

### Data and Statistical Analysis

Line graph representing the optical density (O.D) values of Viral/Antigen load and anti-SARS-CoV-2 antibody titers as a function of time, was constructed with GraphPad Prism version 7.00 (www.graphpad.com). Statistically significant differences between the two methods ‘in house’ ELISA and Euroimmun were evaluated by Pearson’s Chi-squared test with Yates’ continuity correction as appropriate. P<0.05 was considered statistically significant. The samples were examined in duplicates.

### Ethical Statement

Written informed consent was obtained by the patients for the collection and processing of the samples as well as for the publication of this case report. This case report was conducted within the frame of ‘Emblematic action to handle SARS-CoV-2 infection: Epidemiological study in Greece via extensive testing for viral and antibody detection, sequencing of the virome and genetic analysis of the carriers’, which has been approved by the Ethics Committee of Medical School of National Kapodistrian University of Athens (Approval No. 317/12-06-2020).

## Results

Regarding the first assumption of our hypothesis, properly preserved serum from confirmed COVID-19 patients was examined for the presence of viral RNA via RT-PCR. Molecular analysis exhibited a positive result demonstrating that a patient was infected with a previously uncharacterized strain showing 99% similarity with Wuhan-Hu-1 variant **(Suppl. Figure 1A, Suppl. Figure 3A)**, which is in line with evidence supporting its detection in the blood^8–11^. Bioinformatic analysis of this variant revealed the presence of a series of amino acid substitution including the D614G and A879S in the spike (S) protein **(Suppl. Figure 3B)**. Subsequently, to test the second part of our assumption, validation of the produced antibody was carried out in nasopharyngeal swabs, post-mortem material and serum from confirmed COVID-19 patients. The appropriate negative controls were included in all three settings. In swabs, the sensitivity and specificity of the assay reached the values of 96,5% and 99%, respectively (patent application no.:22-0003846810) **(Suppl. Figure 1iii)**. Applying the antibody in immunohistochemical assays, viral particles were detected in archival material from lung tissues of COVID-19 patients^19^. The specificity of the immunohistochemical signal was confirmed by 1) omitting the primary antibody and 2) performing competition with the corresponding spike antigen **(Figure 1B)**. Tissues from a large cohort of non-COVID-19 patients served as negative control^20^. Finally, the ‘in house’ sandwich ELISA assay for antigen detection we developed (for sensitivity and specificity see **Suppl. Figure 1ii)**, identified viral antigens and/or the virus in serum of RT-PCR confirmed COVID-19 patients (antigenemia/viremia), whereas healthy individuals were negative.

Notably, our ‘in house’ sandwich ELISA assay for antigen detection revealed the presence of the virus and/or its constituents in the serum of a patient with remarkably high clinical suspicion of COVID-19 based on his clinical and laboratory findings **(Figure 3A-E, Suppl. Figure 2A)**. In this case, a definite clinical diagnosis was not made possible due to lack of molecular confirmation by RT-PCR in a series of nasopharyngeal (5X) and even a bronchoalveolar lavage (BAL) specimen **(Figure 3A)**. However, post-mortem immunohistochemical analysis of the lung tissue confirmed the viral infection **(Figure 3F, Suppl. Figure 2B)**. Overall, the above workflow demonstrated **a)** the robustness of the antibody produced and **b)** that antigenemia/viremia may take place in a subset of COVID-19 patients.

Given that antigenemia/viremia may occur, we sought to examine whether our working hypothesis stands in the presented case. Monitoring of the kinetics of the two parameters under investigation showed a progressive decline of antigenemia/viremia that was accompanied by a gradual increase of antibody titers resulting in evident seroconversion beyond the expected seroconversion window. In the case of lateral flow testing, seroconversion failed to be detected **(Figure 2B)**.

## Discussion

Immune responses against SARS-CoV-2 have challenged the scientific community worldwide, mainly due to their high inter-individual variability^2^. Heightened immune responses have been associated with adverse clinical outcomes while non-detectable ones, characterize mainly asymptomatic and mild cases^12,21^. The absence of humoral immunity has been attributed to various factors including variability in disease severity and host-related characteristics^22^. With regards to the latter, augmented innate over adaptive immune responses and/or a robust T-cell-mediated viral clearance are proposed as possible explanations^23^. However, this notion was disputed by recent studies demonstrating presence of dominant B-cell over T-cell responses irrespective of disease severity along with longitudinal persistence in memory specific T-as well as B-cells in mild cases^24,25^. This emphasizes the importance of humoral immunity in viral clearance and points towards a different explanation, whilst raising an important question: does the phenomenon of non-responders reflect a true immunobiological event or a limitation of currently available diagnostic tools in precisely portraying the immune landscape?

According to a study comparing different antibody detecting assays, this observation was reproducible by all methods, implying that absence of seroconversion represents a biological rather than a technical issue^22^. To the contrary, serum of presumably non-responders inhibited cell line infection upon culture with viable SARS-CoV-2, as assessed by a neutralization assay, the gold standard for antibody efficiency^12,23^. The latter supports that a technical limitation of the currently available diagnostic tools might be at play^12^.

Our findings suggest that delayed and/or absent kinetics of the antibody reaction is most probably due to the prolonged and increased presence of the virus or its constituents (spike antigen in the present case) in the serum, saturating the antibody response, thus rendering antibody detection feasible only when antigenemia/viremia drops below a certain threshold **(Suppl. Figure 4)**. Similarly to the prolonged shedding seen in nasopharyngeal and gastrointestinal tract secretions, it is likely that such an event could take place in the blood stream, as well^26^. A probable source of antigenemia/viremia could be PANoptosis (Pyroptosis, Apoptosis, Necroptosis), whereby cellular contents are released out of the cell and into the circulation^27^. A similar explanation has been proposed for the presence of viral RNA in the blood, reflecting a wash-out phenomenon from primary sites of infection^9,28^.

The take home message from the current effort is that antigenemia/viremia may affect SARS-CoV-2 antibody tests of seemingly non-responders^23^. In this context, such a phenomenon may lead to delay in or absence of seroconversion (negative result) secondary to immune interference/competition due to immunocomplexes formation **(Suppl. Figure 4)**^15^. It is likely that the ratio between antigens and immunoglobulins -rather the absolute values there of- at a given timepoint during propagation of immune responses determines the result of the immunodiagnostic method. As a result, such a confounding factor should be taken into consideration when interpreting test results. Moreover, it could serve as a partial explanation in discrepancies observed between various diagnostic tools monitoring antibody kinetics, as demonstrated also in the presented case **(Figure 2B)**^23,29^. Therefore, immune response monitoring should be evaluated in longer intervals to better estimate population seroprevalence, thus designing tailored public health strategies (diagnostic algorithms). Such strategies, within the context of vaccine shortage, could potentially include vaccination prioritization of most vulnerable groups harboring little or no antibody protection against SARS-CoV-2 re-infection^30^.

## Data Availability

Not pertinent to this manuscript.

## Acknowledgments

We would like to thank Mr. Dimitris Veroutis for his valuable contribution in performing immunohistochemical analysis in lung tissue specimens, Dr. Panagiota Tsioli in performing RT-PCR in serum of COVID-19-confirmed cases, and Dr. Athanassios Kotsinas, Assistant Professor at the Faculty of Medicine, NKUA, for providing representative NGS results. This work was supported by the: National Public Investment Program of the Ministry of Development and Investment / General Secretariat for Research and Technology, in the framework of the Flagship Initiative to address SARS-CoV-2 (2020ΣE01300001); Horizon 2020 Marie Sklodowska-Curie training program no. 722729 (SYNTRAIN); Welfare Foundation for Social & Cultural Sciences, Athens, Greece (KIKPE); H. Pappas donation; Hellenic Foundation for Research and Innovation (HFRI) grants no. 775 and 3782 and NKUA-SARG grant 70/3/8916.

---

**Supplementary Figure 1. A.** Positive RT-PCR values as detected in the serum of confirmed COVID-19 patients. Amplification of target genes (N1, N2), regions encoding for nucleocapsid protein of SARS-CoV-2, as well as a reporter gene (RNase P) are shown along usage of adequate controls. **B**. Performance characteristics (sensitivity and specificity expressed in percentages) of ‘In house’ assays**: i**. Double Antigen ELISA for antibody detection, **ii**. Sandwich ELISA for antigen detection in serum, and **iii**. Sandwich ELISA for antigen detection in nasopharyngeal swab specimens.

**Supplementary Figure 2. A.** Additional immunohistochemistry pictures depicting presence of SARS-CoV-2 in Type II pneumocytes in the lung of the diseased patient. Staining was performed using secondary antibody 479-G2. **B**. Diagram depicting trends in antigen/viral load and antibody kinetics as a function of time in the same patient. Interestingly, detection of antigen/viral load in the serum was possible via our in house developed ELISA, confirming COVID-19 diagnosis. This is in line with post-mortem immunohistochemical analysis **(Figure 3F, Supplementary Figure 3B)** and in contrast to RT-PCR testing, which was repeatedly negative **(Figure 3A)**.

**Supplementary Figure 3. A.** RNA sequencing of SARS-CoV-2 strain isolated from present case. Upon comparison against Wuhan-Hu-1 complete genome, 99% coverage similarity was observed. **B**. Table listing mutations and their corresponding gene locus within SARS-CoV-2 genome.

**Supplementary Figure 4.** A plausible explanation for interference may be that anti-SARS-CoV-2 antibodies present in the serum may directly cross react to viral antigens and/or virus also found in the serum. Subsequently, the association between anti-SARS-CoV-2 antibodies and viral antigens in the serum will inhibit the cross reaction of anti-SARS-CoV-2 antibodies with the immobilized SARS-CoV-2 antigen immobilized on ELISA microplates. We also suggest that when antigen/viral load decreases, this interference/competitive effect is alleviated, permitting serum anti-SARS-CoV-2 antibodies to cross react to immobilized antigen on the ELISA plate and hence being detected. Competitive ELISA is an established practice in immunoassays where the affinity between an antigen and antibody is tested. We propose that this effect may also occur during serological diagnostic tests for SARS-CoV-2, where antigenemia/viremia observed in COVID-19 will directly interfere with the immunoassay. Presumably, this may also extend to other infectious viral diseases where antigenemia/viremia has been observed.

