## Supplementary material for "SARS-CoV-2 antigenemia/viremia masks seroconversion in a COVID-19 patient": Suppl Figure 1

Supplementary Figure 1

**A**

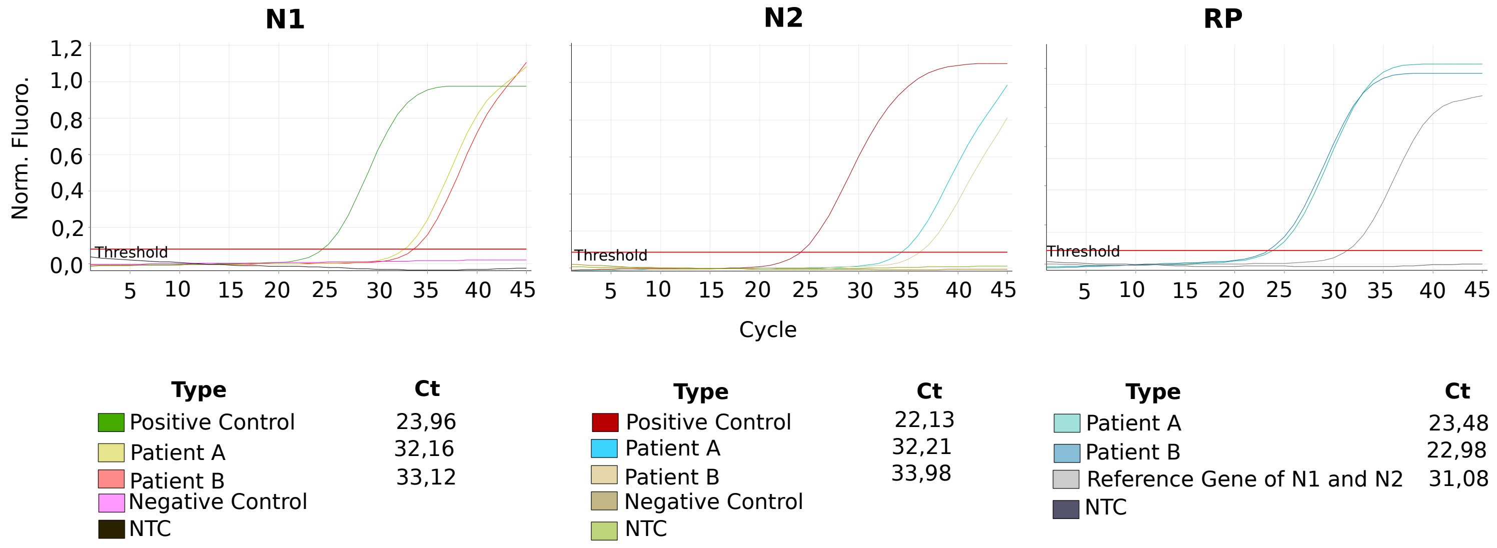


**B**

**i**

Double Antigen ELISA fot Antibody Detection

| | COVID-19 Patients | Non COVID-19 Patients | Total |
|---|---|---|---|
| **Positive Test Result** | 226 | 7 | 233 |
| **Negative Test Result** | 24 | 143 | 167 |
| **Total** | 250 | 150 | 400 |
| | Sensitivity: 90,5% | Specificity: 95% | |

**ii**

Sandwich ELISA for Antigen Detection in Serum

| | COVID-19 Patients | Non COVID-19 Patients | |
|---|---|---|---|
| **Positive Test Result** | 26 | 1 | 27 |
| **Negative Test Result** | 2 | 71 | 73 |
| **Total** | 28 | 72 | 100 |
| | Sensitivity: 93% | Specificity: 99% | |

**iii**

Sandwich ELISA for Antigen Detection in Nasopharyngeal Swabs

| | COVID-19 Patients | Non COVID-19 Patients | Total |
|---|---|---|---|
| **Positive Test Result** | 44 | 1 | 45 |
| **Negative Test Result** | 2 | 123 | 125 |
| **Total** | 46 | 124 | 170 |
| | Sensitivity: 96,5% | Specificity: 99% | |
