## Supplementary figures and images for "SARS-CoV-2 antigenemia/viremia masks seroconversion in a COVID-19 patient"

### Suppl Figure 2

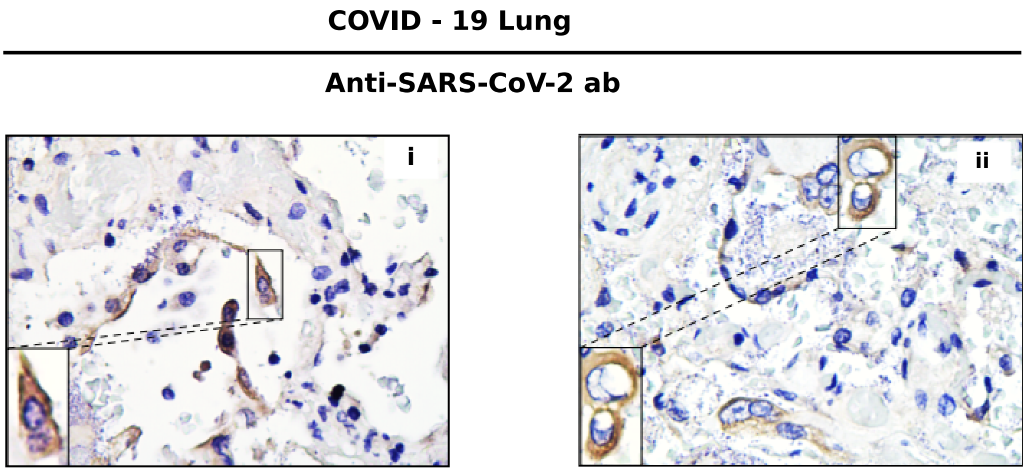

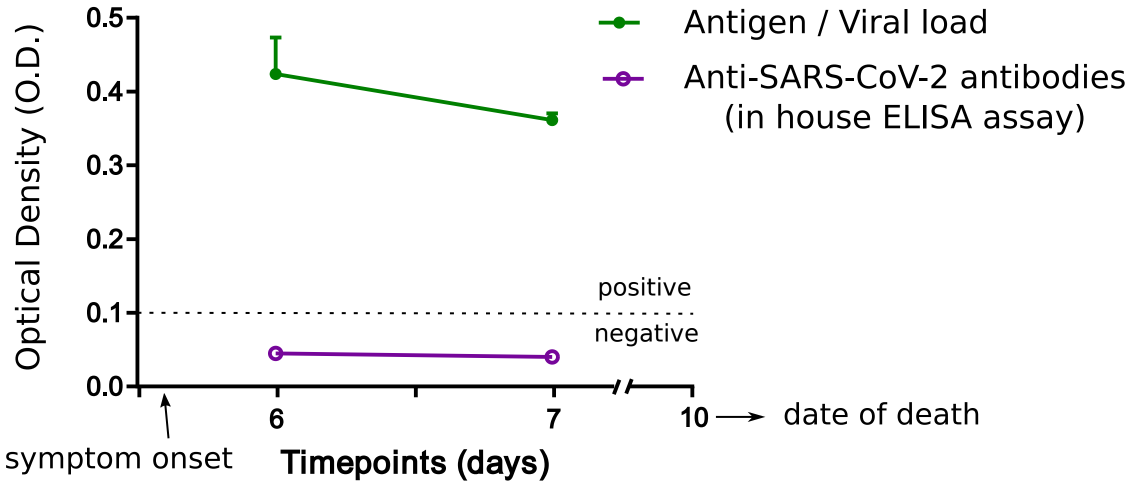

### Suppl Figure 4

Supplementary Figure 4

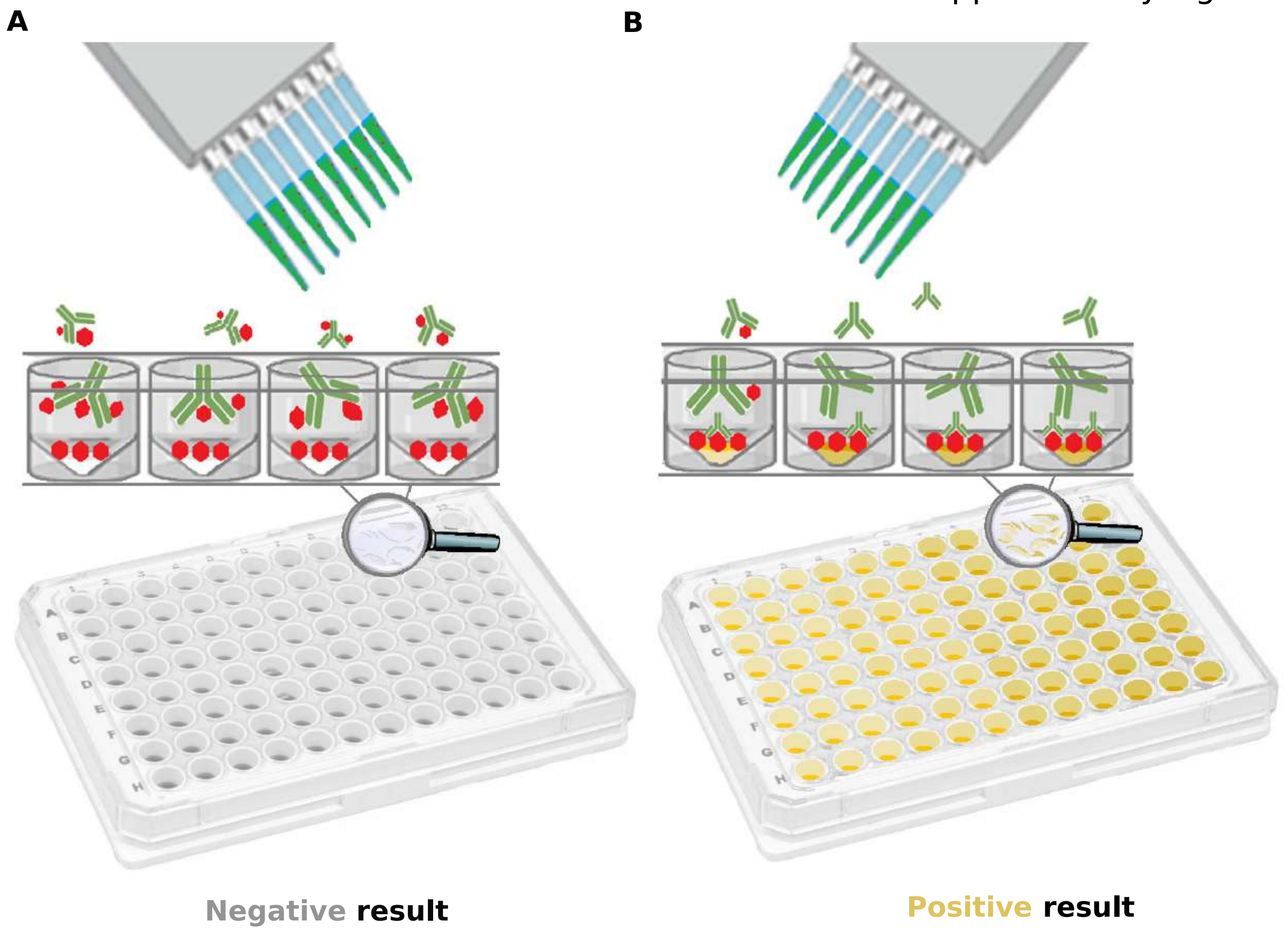
