## Supplementary material for "SARS-CoV-2 antigenemia/viremia masks seroconversion in a COVID-19 patient": Suppl Figure 3

**A**

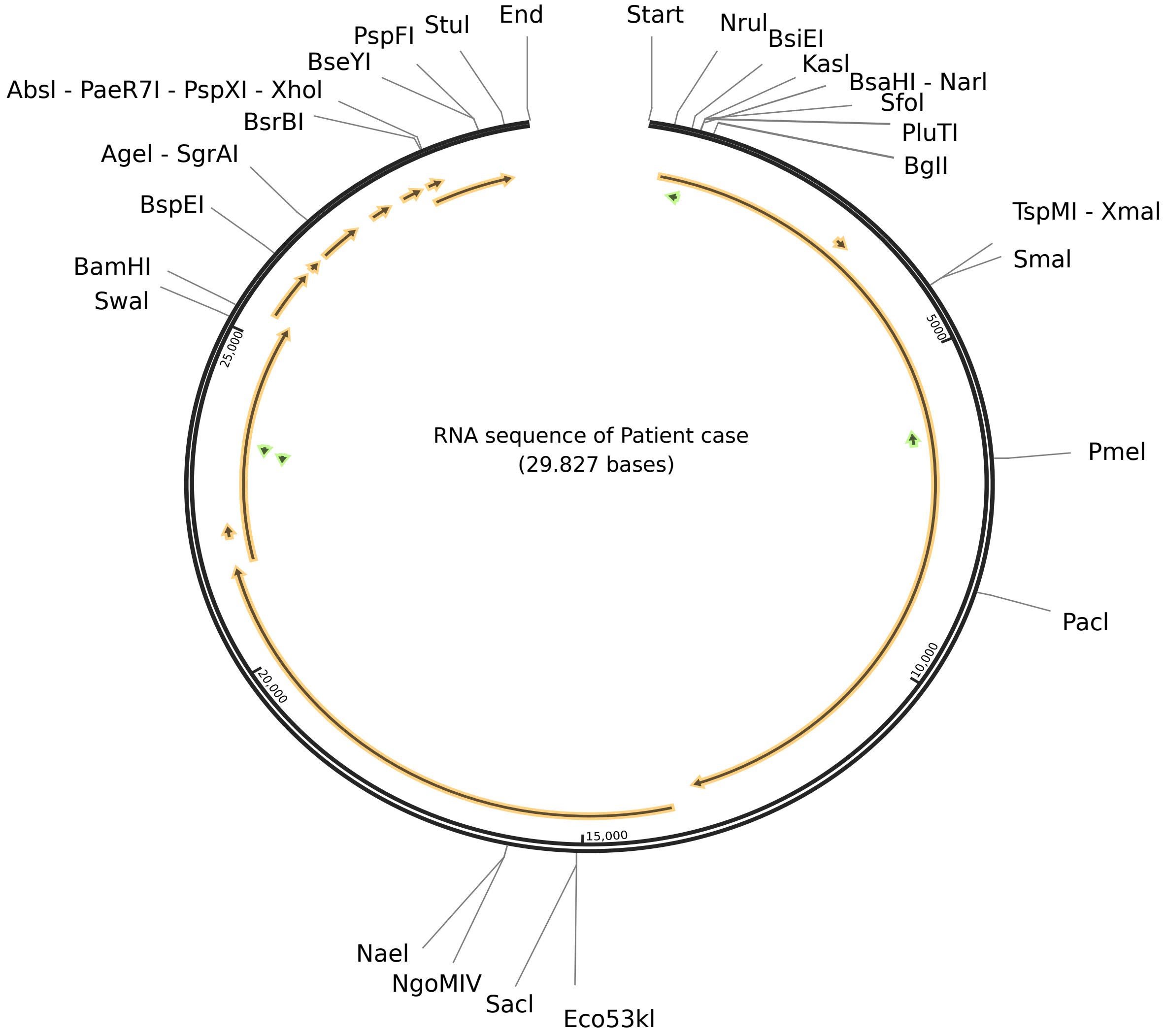


**B**

| List of Mutations | Gene harboring substitutions |
|---|---|
| Q1842R | NSP3 |
| V1897I | NSP3 |
| V34F | NSP8 |
| P323L | NSP12 |
| D614G | Spike |
| A879S | Spike |
| M260K | NS3 |
| T76I | N |
| R203K | N |
| G204R | N |
| D401Y | N |
