## Supplementary material for "SARS-CoV-2 antigenemia/viremia masks seroconversion in a COVID-19 patient": Suppl Materials Methods

### Supplementary Materials and Methods

#### RNA extraction and Real-Time qPCR

A nasopharyngeal swab was collected from a patient with COVID-19-suggesting symptoms and rehydrated in 3 ml of standard viral transport medium (VTM). RNA was isolated from 200µl of VTM and eluted in 50µl using the 740983250 kit (Macherey-Nagel) according to the manufacturer's instruction. Real-Time qPCR was applied as previously described to confirm SARS-CoV-2 positivity[20].

#### Next Generation Sequencing (NGS)

The Ion AmpliSeq Library Kit Plus was used to generate libraries following the manufacturer's instruction, employing the Ion AmpliSeq SARS-CoV-2 RNA custom primers panel (ID: 05280253, Thermo Fisher Scientific). Briefly, library preparation steps involved reverse transcription of RNA using the SuperScript VILO cDNA synthesis kit (Thermo Fisher Scientific), 17-19 cycles of PCR amplification, adapter ligation, library purification using the AgencourtAMPure XP (Beckman Coulter), and library quantification using Qubit Fluorometer high-sensitivity kit. Ion 530 Chips were prepared using Ion Chef and NGS reactions were run on an Ion GeneStudio S5, ion torrent sequencer (Thermo Fisher Scientific).

#### Bioinformatics

The SARS-CoV-2 Wuhan-Hu-1 strain complete genome was used as reference for alignment. Both, AmpliSeq alignments and quality controls were performed using the Torrent Server of

Ion Torrent S5 sequencer employing default settings. Aligned reads served for both reference-guided assembly and variant calling. Assembly was performed using the Iterative Refinement Meta-Assembler (IRMA v0.6.1) that produced a consensus sequence (per sample) using a >50% cut-off for calling single nucleotide polymorphism.
